# TeleAutoHINTS: A Virtual or Augmented Reality (VR/AR) System for Automated Tele-Neurologic Evaluation of Acute Vertigo

**DOI:** 10.1101/2025.03.14.25323774

**Authors:** Haochen Wei, Justin Bosley, Edward Kuwera, Peter Kazanzides, Kemar E. Green

## Abstract

Annually, several million patients in the United States visit the emergency room (ER) with symptoms of vertigo or dizziness. Rapidly distinguishing between benign causes, such as inner ear disease, and more severe conditions, like strokes, necessitates performing and interpreting a three-step bedside head and eye movement assessment called HINTS (head impulse, nystagmus, and test of skew). This test is more accurate than state-of-the-art brain imaging, especially early in the disease’s course when there is a limited window for safely conducting lifesaving stroke interventions. A significant barrier to its widespread adoption is the shortage of experts trained to safely perform and interpret this test in the ER. This highlights the need for automated remote assessments. In response, we developed TeleAutoHINTS as a two-part solution: (1) a head-mounted display-based head and eye tracking platform, using Microsoft Hololens2, for automated tele-sensing and (2) an interconnected interface for real-time data visualization and analysis. We tested TeleAutoHINTS on three subjects to assess the feasibility of automated testing and evaluate the head and eye movement recordings. Preliminary results suggest that a head-mounted display-based remote self-assessment platform for acute vertigo diagnosis is technically feasible.

## 1 Introduction

Dizziness/vertigo affects millions of patients in the U.S. annually [1], [2]. The causes of dizziness and vertigo can be neurologic or nonneurologic [3]. The neurologic causes can be accurately delineated using key components of medical history, as well as eye movement and head position assessments [4]. In acute settings, distinguishing rapidly and accurately between benign inner ear diseases and potentially debilitating (or fatal) neurologic injuries (usually strokes) in patients experiencing a sudden onset of constant dizziness (or AVS - acute vestibular syndrome) has garnered significant attention in neuro-otology and neuro-ophthalmology [5]. High-resolution brain imaging often fails to accurately differentiate these groups during the critical window for urgent interventions [6]. The inadequacies of brain imaging have contributed to increased healthcare spending on unnecessary tests and increased patient disability [2]. As a result, experts developed a three-step testing battery known as HINTS (Head Impulse, Nystagmus, and Test of Skew). This test is grounded in the physiology of the neural circuits connecting the inner ear’s rotation and gravity sensors to the brain [4,7,8]. It evaluates changes in eye movement based on gaze positions, the vertical alignment of the eyes within the orbit, and subtle eye movement reactions to high velocity/low amplitude operator-induced head rotations. In the appropriate time frame for patients with AVS, HINTS has proven to be more sensitive and specific than a brain MRI in differentiating strokes from vestibular neuritis (inner ear disease) [6].

Although the HINTS test exists and data support its accuracy in identifying strokes in acute vertigo patients, its widespread adoption faces challenges. The primary challenges are the availability of vestibular neurology experts to interpret or administer the test and the scarcity of skilled emergency room providers trained in the technique [9,10,11]. This dearth of skilled individuals has prompted a reimagining of the HINTS assessment, leading to the popular use of portable video-oculography systems [12] and various telemedicine solutions [13,14,15,16]. Yet, none of these fully address the root problem. Consequently, we propose TeleAutoHINTS, a virtual/augmented reality solution for the automated tele-assessment of acute vertigo patients. We also conduct preliminary patient testing to evaluate the feasibility of this innovative approach. TeleAutoHINTS (Fig. 1) has the potential to reduce the need for skilled operators and amplify the services of the limited expert group to health centers both nationally and internationally. The concept of remote sensing of head position and eye movement with AR and VR guidance has further clinical implications beyond assessing acute vertigo.

**Figure 1:**
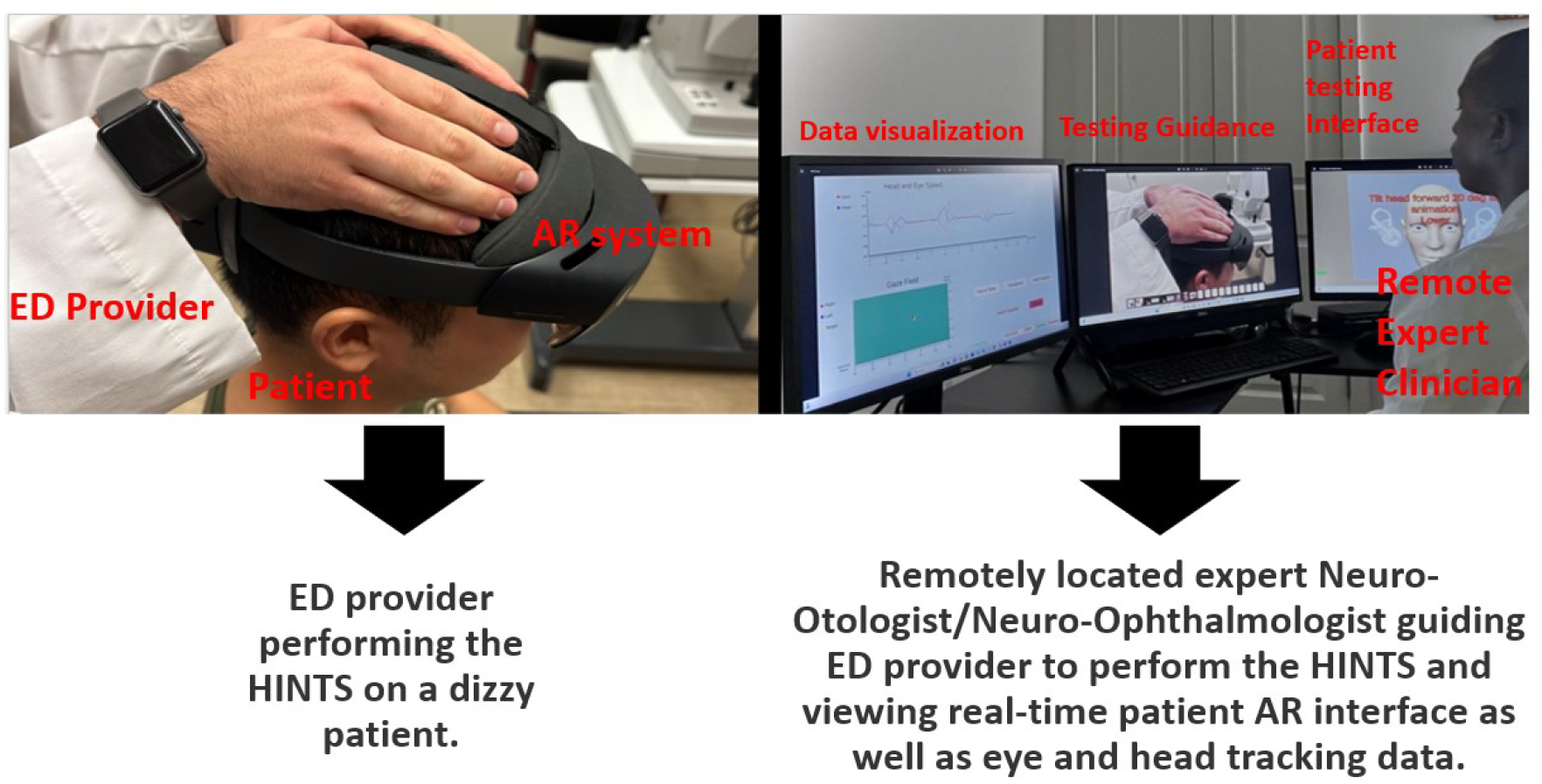
Concept of TeleAutoHINTS: A Virtual or Augmented Reality System for Automated Tele-Assessment of Acute Vertigo. ED provider - Emergency Room Medical Personnel (e.g., physician, physician assistant, nurse practitioners, etc.).

## 2 VR/AR Applications in Eye Movement and Vestibu-lar Neurology

Virtual and augmented reality eye tracking is an emerging area in neurology, ophthalmology, and other related fields, creating novel opportunities for research and clinical applications [17,18,19]. In recent years, researchers have begun to compile open-source VR eye tracking datasets for various clinical and non-clinical use-cases [20,21], facilitating cross-institutional AR/VR eye tracking research. In clinical neurology and ophthalmology, VR/AR eye tracking solutions have been implemented for the measurement of eye alignment [22,23,24,25,26], other eye movement abnormalities [27,28,29], and the treatment of eye movement disorders [30]. In vestibular neurology, VR/AR has been utilized to quantify vestibular function [22,31], evaluate balance in vestibular migraine patients [32], treat benign paroxysmal positional vertigo [33], and provide vestibular rehabilitation for various causes of dizziness, vertigo, and imbalance [34,35,36,37,38,39,40,41,42]. To date, no one has leveraged AR or VR head-mounted displays for the automated tele-assessment of acute vertiginous patients.

## 3 Materials and Methods

The TeleAutoHINTS system comprises two parts: the head-mounted display-based testing platform (Patient Testing Interface [PTI]), designed to guide patients through the test, and the desktop visualization and validation platform (Medical Provider Interface [MPI]). The MPI allows providers to switch between automatic data collection (patient-initiated) and quasi-automatic data collection (provider-initiated). It also offers an interface for real-time monitoring and post-test data analysis. Both interfaces were created using Unity3D (version 2021.3.28f1).

### 3.1 Patient Testing Interface (PTI)

We implemented PTI on Microsoft HoloLens 2, which has a sampling rate of 60Hz and a resolution of 2048 × 1080 pixels. The horizontal and vertical fields of view are 43° and 29° respectively, which allows for the optimal assessment of eccentric gaze eye position data [8]. We leveraged the HoloLens’ built-in eye tracker to collect gaze data. Microsoft did not disclose their gaze tracking accuracy and precision, but Microsoft’s developer website provides a rough estimate of 1.5°. We obtained the gaze data stream from each eye using the API provided by Microsoft at 60 fps. Due to Microsoft’s restrictions, a consent and calibration program must be conducted before collecting gaze data. This means that all our recorded gaze data has been calibrated using HoloLens 2’s internal calibration program. We used the built-in research mode to obtain gyro data, which helped in calculating head position. The HoloLens did not provide any raw eye movement video data. We used Microsoft’s Mixed Reality Toolkit (MRTK), which provides a toolkit for integration between HoloLens and Unity3D (Version 2021.3.28f1). We incorporated the Microsoft-provided Extended Gaze Tracking API and the HoloLens2-ResearchMode-Unity Package [70].

Because HoloLens 2 is an optical see-through head-mounted display, our application can be considered augmented reality (AR). However, it could alternatively be implemented in virtual reality (VR), assuming the availability of sufficiently accurate gaze tracking on this platform.

The PTI user interface, shown on the left in Figs. 2-4, includes several components: the visual target, an “OK” button, a “Cancel” button, instructional display text, and the connection status. Recognizing the range of technological skills among potential patients, we designed the user interface to be as simple as possible. Upon launching the program, the only visible option is an “OK” button to initiate the test. The HINTS test consists of three subtests: head impulse, nystagmus, and the test of skew, described in the following subsections. Consequently, patients must complete all subtests consecutively. Between each subtest, there is a patient-controlled resting period when the “OK” button is activated, allowing patients to proceed when ready. The “Cancel” button is available throughout the test. If pressed during a subtest, that specific subtest stops, and the ongoing test is marked incomplete. The patient can then press “OK” to restart the interrupted test. Pressing the “Cancel” button when a subtest is not active terminates the entire test session, returning the system to the phase before the first subtest. If no gaze data is detected, a warning is displayed, alerting the user to a potential calibration problem with their device. When the MPI is connected to the PTI and a test is initialized, the medical provider takes full control of the test procedure, including the ability to choose, start, or stop subtests. Once a provider takes over, patients cannot initiate test actions but can still halt or exit the PTI.

**Figure 2:**
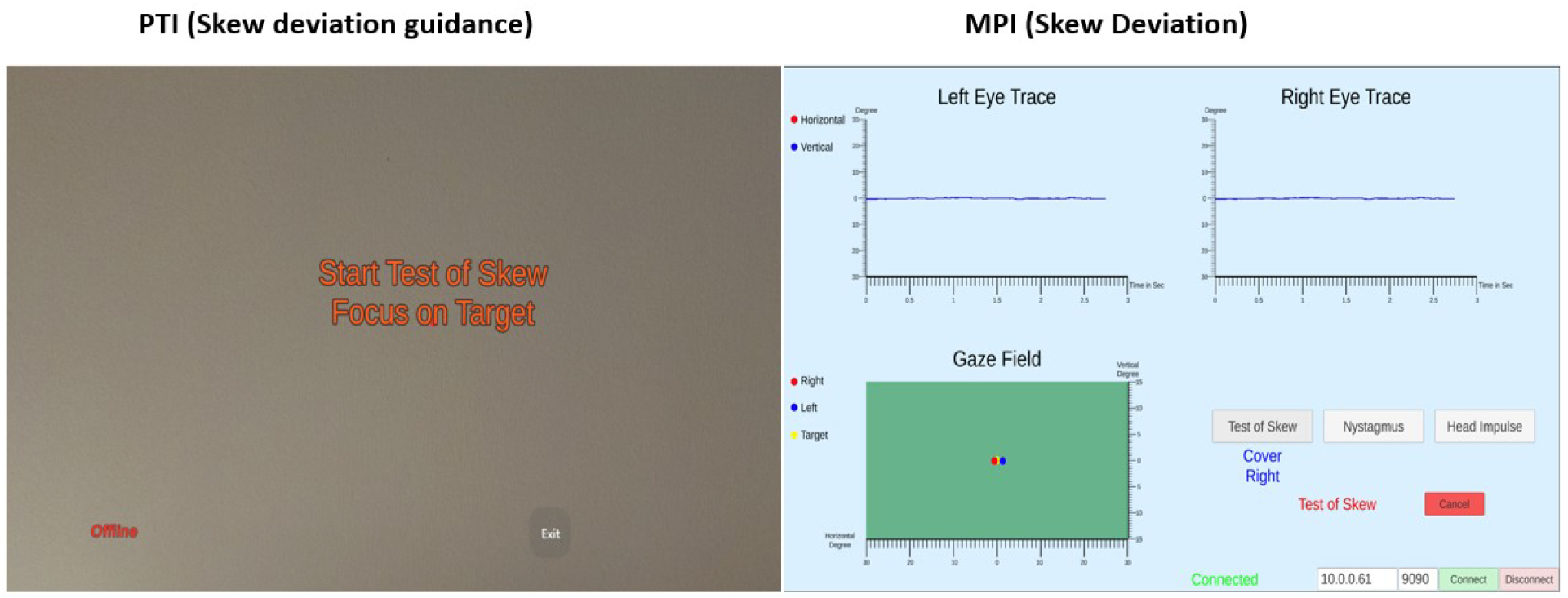
Skew deviation guidance in the PTI and corresponding vertical gaze position in the MPI.

#### 3.1.1 Test of Skew

Traditionally, skew deviation is assessed by stabilizing the head and focusing on a single fixed object while alternating eye coverage. However, conducting this test solo is impractical for the patient. Thus, we simulate the cross-covering technique by displaying the target object in only one eye, eliminating the need for physical movement. Once the test starts, the PTI instructs the patient to focus on the target using displayed instructional text, as shown in Fig. 2. The PTI then alternates the displayed eye, recording eye gaze data for test execution. The entire test duration is 15 seconds, including a 3-second initialization stage where both eyes can see the target. Data recorded during this stage is not analyzed as it is meant for patient adaptation. This is succeeded by four cross-covering stages, each 3 seconds long, where the target alternately appears in one eye.

#### 3.1.2 Nystagmus Test

The Nystagmus test requires data collection both with and without a fixation target. The device offers guided instructions, directing patients to either fixate on a target or look away, across various eye positions. The entire test duration is 100 seconds. The first 10 seconds involve looking straight with the target displayed, followed by looking left 20 degrees for 10 seconds with the target displayed, then straight for 10 seconds, right for 10 seconds (both with the target displayed), and straight again with the target displayed for another 10 seconds. This concludes the fixation nystagmus test. The subsequent non-fixation nystagmus test sequence is: look straight for 10 seconds without target, left for 10 seconds without target, straight for 10 seconds without target, right for 10 seconds without target, and finally, straight-ahead for 10 seconds without any target. To demonstrate the system’s nystagmus detection capability, we simulated physiological horizontal nystagmus using an optokinetic (OKN) stimulus moving horizontally at 1.5 Hz as shown in Video 2 and Fig. 3. Throughout, patients are instructed to maintain a straight-ahead gaze. The test lasts 23 seconds, but the initial 3 seconds are disregarded as patient adaptation time.

**Figure 3:**
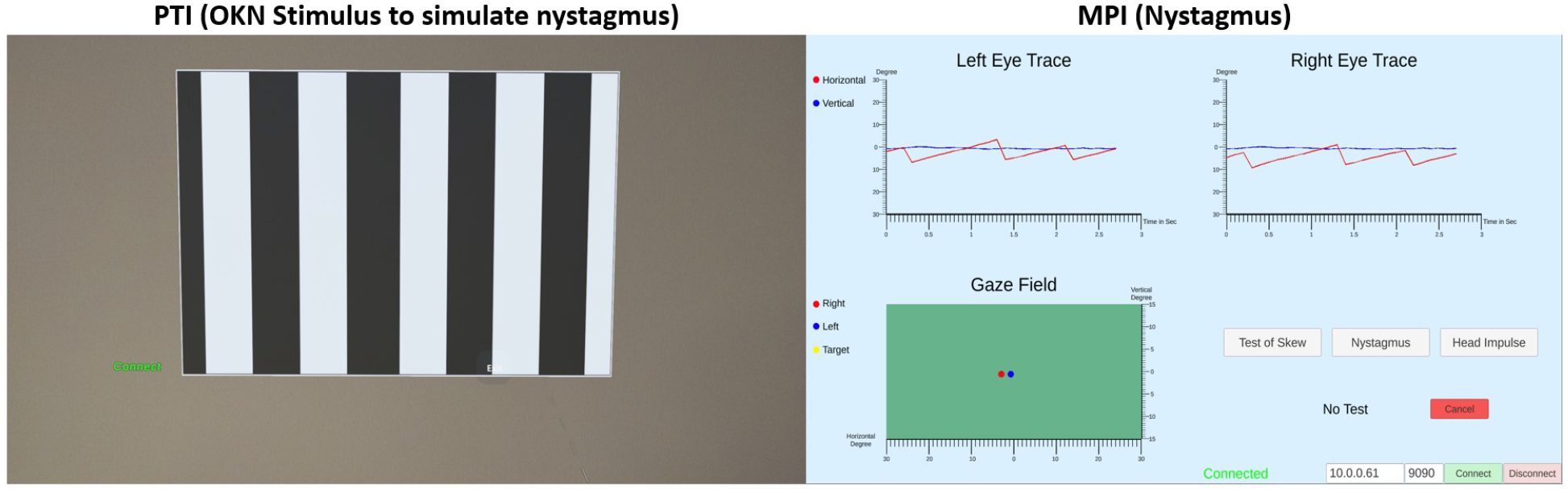
Optokinetic stimulus in the PTI and corresponding left-beating nystagmus (slow-phase to the right and fast-phase to the left) in the MPI.

#### 3.1.3 Head Impulse Test

The Head Impulse test can be conducted with (passive) or without (active) the aid of an operator. Current research does not widely support active head-impulse due to potential inaccuracies[43], [44], and safety concerns. While anecdotal evidence suggests its utility for remote acute vertigo diagnosis during the pandemic[13], more research is needed to affirm its clinical value. Therefore, we included a medical-provider-directed head impulse. In this approach, a remote medical professional instructs on-site care providers to perform passive head impulses, providing real-time feedback, as shown on the left in Fig. 4. On the PTI end, the patient is instructed to tilt their head forward by 20 degrees, ensuring they visually fixate on a target set at 0 degrees, as shown in Fig. 4. From this position, unpredictable head impulses in the yaw plane can be executed with velocities of ≥ 120 degrees/second and amplitudes of ≤ 10 degrees to either side of the visual target, allowing a latency of ≥ 1 second between each impulse. The target remains fixed at 0 degrees during these quick head movements. The device continuously monitors head speed and starts recording for 500 ms when the current head speed exceeds 50 degrees/sec. Head impulse results with a peak velocity of <120 degrees/sec are excluded during the data analysis phase. Data is streamed to the MPI regardless of the recording status.

**Figure 4:**
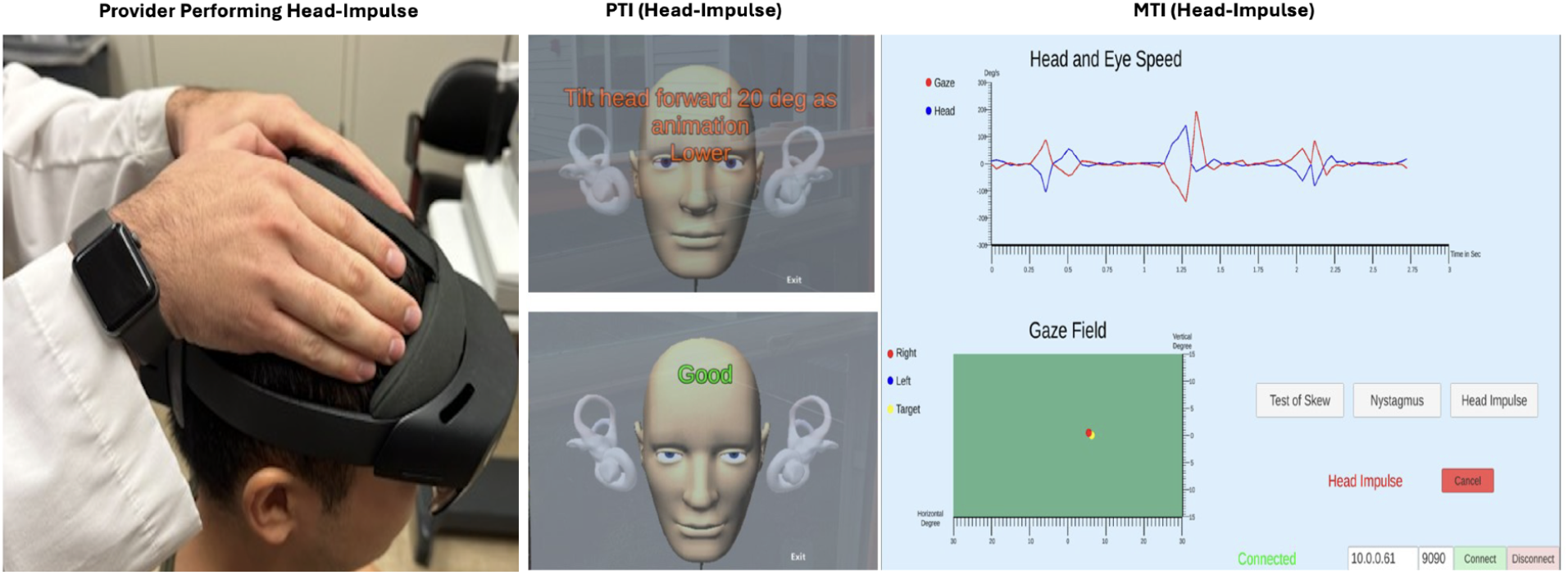
Setup for head-impulse test with PTI guiding head positioning and MPI showing accepted head-impulse data.

### 3.2 Medical Provider Interface (MPI)

This system is divided into two primary components: the User Interface and Visualization, and the Communication Portal. The User Interface and Visualization system is composed of three primary elements: Gaze Visualization (GV), Connection Control Panel (CCP), and Test Control Panel (TCP), as shown on the right in Figs. 2-4 and Video 1-2.

The GV displays the pupil position data from both eyes for the test of skew (vertical only) and nystagmus tests (horizontal and vertical). For the head impulse test, it displays head and eye velocity-time graphs that show the head thrust and its corresponding opposite direction eye movement response. This component retains and presents gaze data from the past three seconds, enabling remote medical providers to verify the testing. It also tracks the real-time gaze field, illustrating the eye position in reference to the target position throughout the recording. For the skew deviation test, there is a text prompt that indicates in real-time which eye is covered or uncovered.

The CCP is utilized to establish and manage the connection with the PTI and to show the connection status. This is shown in the lower right corner of the MPI interface (see Figs. 2-4

The TCP permits medical providers to initiate, repeat, or cancel any test during the connection. Once a doctor assumes control of the test, the patient loses the ability to influence it. The test’s status is also displayed within this panel, which is also in the lower right corner of the MPI interface, just above the CCP.

The Communication Portal operates on a TCP socket and interfaces with the PTI. Serving as a client, it receives head and gaze data streams for visualization and validation and sends commands to oversee the testing process. There exists latency between the PTI and MPI. Although we did not comprehensively examine this latency – as it can fluctuate based on the network environment – it is worth noting that, since all our data collection and testing procedures operate locally on the PTI, and all the raw data is transmitted afterward, latency is not a paramount concern for our system.

## 4 User Study

We conducted a preliminary study to determine if our TeleAuto-HINTS system could collect meaningful data from patients.

### 4.1 Participants

All participants (n=3) were adults (≥ 18 years old) undergoing inperson video-oculography (VOG) and video head-impulse (vHIT) testing due to complaints related to various neurological illnesses. All participants consented to participate in the research. The study received approval from the institutional review board. All subjects underwent their clinical VOG and vHIT tests before using TeleAu-toHINTS. As described above, TeleAutoHINTS comprises three subtests (Video 1): skew deviation, nystagmus, and head impulse.

### 4.2 Head and Gaze Data Analysis

Post-session data analysis and visualization were performed using Matlab. The extended gaze tracking API and HoloLens 2 provided timestamps. Gaze and Head Velocity were calculated based on the eye and head position, divided by the time frame difference. All data were transmitted at 60 fps from the PTI to MPI. Data from the nystagmus test and test of skew did not undergo any post-processing. HoloLens 2 also reported the eye position in 3D space, which could be used to compensate for eye convergence. However, for our testing purposes, only raw data was collected and analyzed. The Head Impulse test data was interpolated using spline interpolation built in Matlab and then plotted at 240Hz. Inappropriate data points were identified and excluded before plotting. This includes instances like blinking during the head impulse, which leads to gaze frame loss, and non-optimal head impulses that are too slow (below 120 degrees/s).

### 4.3 TeleAutoHINTS Results

As shown in Fig.5, the TeleAutoHINTS test results from all three subjects demonstrate that the system can produce high-quality head and eye traces that mimic physiologic eye movement responses to head impulses with the appropriate amplitude, velocity, and duration. The head velocity data from the head impulse exceeds 120 degrees per second for all subjects, aligning with clinical standards. This is crucial since low-velocity head impulses can yield inaccurate vestibular function information. The simulated nystagmus waveforms are robust, closely resembling the typical jerk nystagmus waveform morphology with an initiating slow phase followed by a fast phase in the opposite direction. This alternating pattern of slow and fast phases (or a beat) allows medical providers to assess: 1) The direction of the nystagmus (named for the direction of the fast phase); 2) The side of the brain or inner ear affected (indicated by the direction of the slow phase), and 3) The slow-phase velocity and amplitude of the nystagmus, which can aid in differentiating between brain and inner ear nystagmus in both fixation and fixation-removed scenarios. Lastly, the test of skew measures the degree of ocular misalignment in terms of vertical eye rotation.

**Figure 5:**
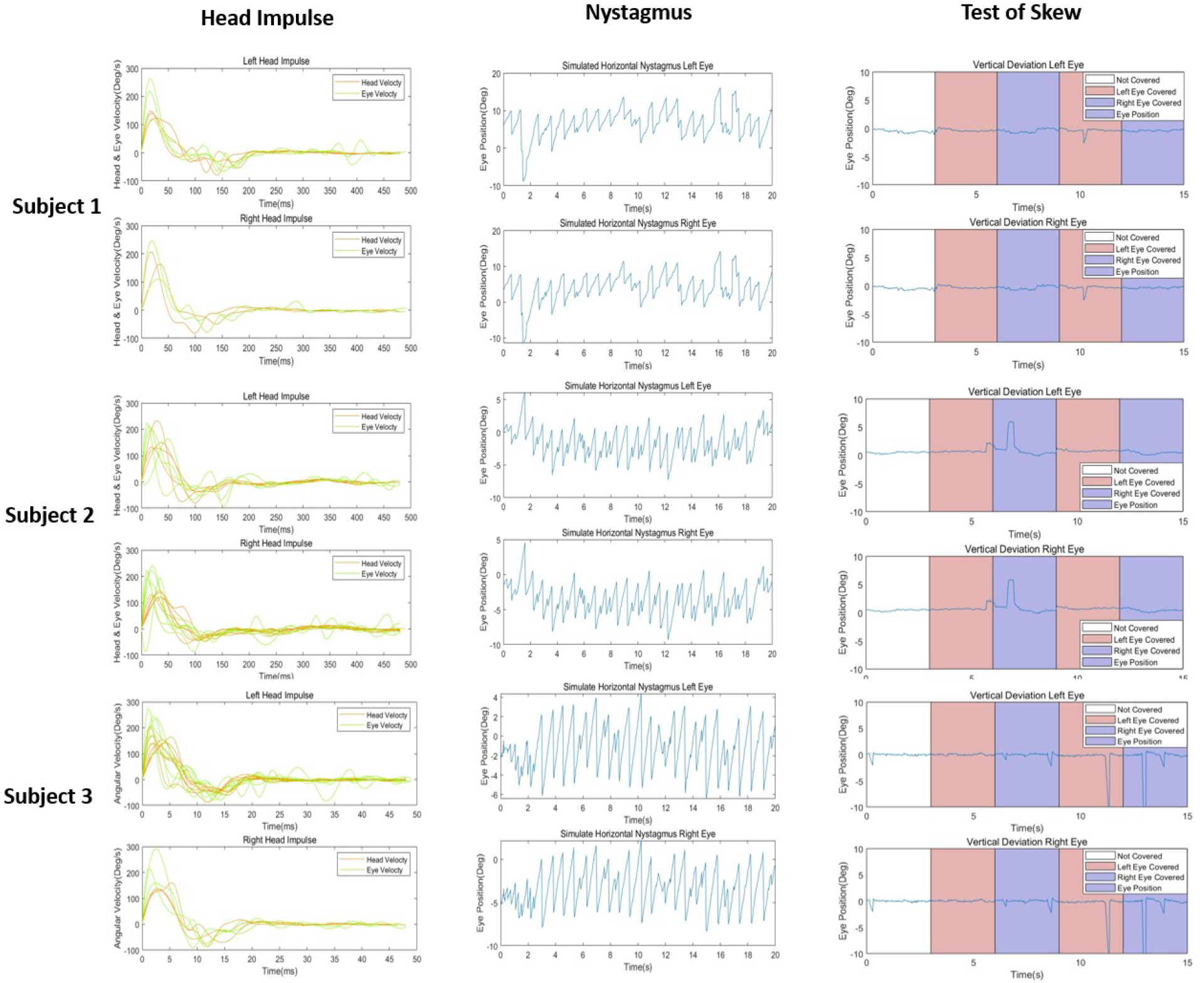
HINTS testing results for the three subjects. All subjects had normal head-impulse test defined as an eye to head velocity ratio of ≥ 0.7. All 3 nystagmus traces demonstrate fast-phases to the right (upward defection) or right-beating nystagmus. Subject 3 had the largest amplitude of all three. All three tests of skews were normal with artifacts (blinks) in all three mimicking vertical deviation of the eyes.

## 5 Discussion

At its core, HINTS [4] focuses on recognizing nuanced eye movement variations due to disturbances in the neural pathway, which spans from the inner ear to brainstem structures[7,8,43]. These structures regulate eye movement and the orientation of the head with respect to the earth’s vertical axis. The HINTS test profile for an acute brainstem/cerebellar stroke includes a normal head impulse test, direction-changing nystagmus, and the presence of skew deviation (vertical misalignment of the eyes). This profile is more sensitive and specific than an MRI of the brain in the first 48 hours. Despite its known limitations[9,11,45–50], the HINTS and its extended version, HINTS “plus” (which includes a hearing loss component) [45], have consistently demonstrated their value in assessing patients with AVS in emergency settings.

Given the scarcity of VOG and HINTS-expert professionals, there is a clear demand for a tele-automated system. Regular training for emergency room staff is not always feasible[51,52]. Our solution, TeleAutoHINTS, addresses this need by eliminating the continuous training requirement, offering a remote self-directed data collection method via a VR/AR headset. We also showcased the feasibility of autonomously collecting high-quality eye movement data using AR/VR guidance. Initially, TeleAutoHINTS only automates the tele-sensing of certain HINTS components (nystagmus and skew test). However, its potential to reliably gather data on operator-dependent head impulses has been evident. As the system evolves, we aim to integrate three head-impulse testing methods: clinician-directed (passive), robotic arm-guided (passive)[53], and operator-independent (active)[13,15]. The last method will be included after its validation. Moreover, alternative techniques that avoid actual head thrusts might be integrated as proxies for head rotation[54] to enhance telemedicine systems.

TeleAutoHINTS also excels in collecting quantitative data, which experts[55] can then remotely evaluate. Recent academic efforts have explored integrating artificial intelligence (AI) into various HINTS aspects[49], [56,57,58,59,60]. Such progress sets the stage for future versions of our system to leverage AI, particularly beneficial for facilities without in-house expertise. Incorporating AR/VR technologies ensures easier adoption of supplementary methods like video-ocular counter roll (vOCR) [61], hearing tests[45], fundus photography[47][49], gait analysis[50], and others[61], enriching the Tele-HINTS process. This advantage mainly stems from AR/VR’s superior immersion, adaptability, and compatibility[62,63,64,65,66] compared to HINTS methods based on smartphones[67,68,69].

The current system has two primary limitations. First, the absence of raw eye movement videos in the HoloLens hampers clinicians from validating ocular motor responses accurately. This feature becomes essential when eye tracking capabilities falter, necessitating direct video inspection for diagnosis. We aim to rectify this by deploying our system in a device offering raw eye tracking video outputs. The second limitation is that the system is not integrated with existing teleconferencing platforms. In response, we plan to design an interface compatible with prevalent telemedicine streaming platforms.

## 6 Conclusion

Traditional HINTS methodologies, while clinically invaluable, face significant challenges, particularly in emergency settings due to the lack of specialized knowledge and the absence of VOG tools. We have demonstrated that it is feasible to automatically collect components of the HINTS using our AR/VR-powered TeleAutoHINTS system. TeleAutoHINTS, harnessing the potential of AR/VR, addresses these challenges with its telemedicine-enabled self-directed protocols, modernizing essential HINTS components. As technology advances, the role of AI is set to expand, providing more streamlined analysis, especially in facilities grappling with expert shortages. By potentially integrating a wide range of diagnostic methods, TeleAuto-HINTS stands to revolutionize acute vestibular syndrome evaluations in emergency situations. Furthermore, this system is poised to become an invaluable tool for specialists like neuro-ophthalmologists, neuro-otologists, strabismus surgeons, and others involved in clinical roles or research within the realms of vestibular neurology and eye movement disorders.

## Data Availability

All data produced in the present study are available upon reasonable request to the authors.

